# Investigating the Readability, Visual Design, and Quality of Online Written Pharmacogenomics Health Information for Health Consumers in Australia

**DOI:** 10.64898/2026.05.27.26354169

**Authors:** Matthew J. Giblett, Yousef Babikian, Dillensinh J. Jhala, Sarah E. Medland

**Affiliations:** Psychiatric Genetics, Brain and Mental Health, QIMR Berghofer, Brisbane, QLD, Australia; School of Psychology, University of Queensland, Brisbane, QLD, Australia; School of Psychology and Counselling, Queensland University of Technology, Brisbane, QLD, Australia

**Keywords:** pharmacogenomics, health information, health on the internet, health literacy, personalised medicine, readability

## Abstract

Pharmacogenomics (PGx) offers a pathway towards personalised medicine, which relies on health consumer involvement in making informed decisions. As consumers increasingly seek health information online, high-quality digital resources are essential to support informed consent and shared decision making. The complexity of PGx and widespread limitations in health literacy raise concerns about whether existing consumer-facing online PGx resources are understandable and sufficiently comprehensive. This study evaluates the readability, visual design, and informational quality of publicly available online written PGx health information. Twenty-three webpages met inclusion criteria. The mean readability corresponded to approximately 15 years of formal education (university level), substantially exceeding the Australian Government’s recommended Year 7 reading level for public health materials. Informational quality was generally low, with most webpages being rated as poor or very poor. In contrast, visual design quality was relatively strong, with webpages achieving on average around three-quarters of the criteria. Although the visual presentation of PGx webpages is generally professional, their high reading difficulty and limited discussion of treatment choices and uncertainties reduce their usefulness for health consumer education. Improving readability, clearly communicating risks and limitations, and incorporating decision-support features may enhance the ability of online resources to support informed consent and shared decision making.

## Background

Pharmacogenomics (PGx) is the study of peoples’ genetic variations affecting metabolic enzymes, drug receptors, and drug transporters, which cause increased risks of adverse drug reactions or decreased drug efficacy (Wake et al., 2019). Adverse drug reactions are negative consequences resulting from medication use in accordance with proper therapeutic use and dosage, or the potentially preventable morbidities and mortality associated with the otherwise appropriate use of a medication (Nebeker et al., 2004; Wester et al., 2008). Currently, medication prescription often follows guidelines which assume “one-size-fits-all” or “trial and error” methods, whereby health consumers cycle through various treatments until an effective match is found (J. T. Y. Cheng et al., 2025; Suppiah et al., 2018). PGx presents a pathway towards personalised medicine, where the aim is to deliver more patient-centred care guided by personal characteristics including genetic, lifestyle, environmental, and other biological considerations (Sadee et al., 2023).

Tailoring medication selection and dosage to consumers’ specific features surpasses the one-size-fits-all model by maximising efficacy and minimising toxicity for the individual (Giacomini et al., 2012; Ibrahim, 2025). PGx offers several significant benefits to the healthcare system. In Australia, it is estimated that it could save A$2.5 billion to A$6.2 billion over a 5-year period from trial-and-error wastage (Suppiah et al., 2018). It also provides a pivotal opportunity to predict and prevent adverse drug reactions, thus easing healthcare resource utilisation, improving patient outcomes, and possibly saving another A$1.0 billion to A$1.6 billion a year (Bi et al., 2025; Chenchula et al., 2024; Suppiah et al., 2018). In an Australian First Nations context, some health inequities are tied to expecting one-size-fits-all to work for all Indigenous groups, despite the existence of ethnically distinct PGx variants between Indigenous communities which should guide personalised medicine for individuals within those communities (Nasir et al., 2022; Samarasinghe et al., 2023). However, current research on PGx depends on samples overrepresented by those of European ancestry, severely restricting First Nations consumers’ access to reliable personalised medicine (Lewis et al., 2024; Pirmohamed, 2023). First Nations consumers seeking education on PGx and personalised medicine deserve to be adequately informed of both the potentials and limitations of the field to avoid misplacing their hopes and empower them to engage in shared healthcare decision making.

The personalisation of health care relies on shared decision making. It is built upon collaborative partnerships in which consumers and clinicians determine the best options by weighing clinical findings alongside the consumer’s values, preferences, expectations, and socioeconomic contexts (Burke et al., 2014; Salari & Larijani, 2017; Shickh et al., 2023). Consumer involvement in making informed health decisions is a fundamental right, central to safe and high quality health care, with shared decision making representing the highest standard of informed consent (Australian Commission on Safety and Quality in Health Care, 2021; Hoffmann et al., 2025).

Informed consent requires three fundamental criteria: the individual must be competent, adequately informed, and not coerced into making a decision (Cocanour, 2017). Ensuring effective shared decision making and informed consent requires access to trustworthy, accurate, and accessible information and expertise both within and between clinical encounters (Shickh et al., 2023). As clinical consultation time is often limited, consumers increasingly turn to digital resources for health-related information. Nearly all Australians continuously access the internet on a personal device at home (Australian Communications and Media Authority, 2024), with around three quarters of adults using the internet to find health-related information at some point over a 12-month period (Research Australia, 2017, 2019). For general practice patients, nearly a third sought health information online at some point over a one-month period, of which a quarter of those pursued medication-related information (Wong et al., 2014). Internationally, a 2022 web-based survey of 1,092 adults in Poland found that nearly two-thirds of participants searched the internet for health information, with the most common search being for information on medications and their effects (Płaciszewski et al., 2022). The prevalence of these search behaviours highlights the internet’s role as a primary source of health information—especially related to medications—between clinical encounters.

The Australian Government’s *Style Manual* (Australian Government, 2024) recommends that online sources should be written at a lower secondary level (year 7, or between 12 and 14 years old). This is because around 44% of Australian adults read at literacy levels 1–2, or between pre-primary school to year 10 (Australian Bureau of Statistics, 2013). Australian health consumers also have low health literacy levels (Choudhry et al., 2019), which acts as an additional barrier to informed consent and shared decision making (Hoque, 2024; Muscat et al., 2021; Ng, 2024). Low health literacy is also associated with greater risk for adverse drug reactions (Gupta et al., 2020). Despite widespread low literacy levels, previous research has found that the average online health webpage in Australia is three to five grades higher than the Government’s recommended level (C. Cheng & Dunn, 2015). This indicates that health webpages may not be adequately considering the literacy levels of the general public.

PGx information is complex and can be difficult for both consumers and healthcare providers to understand (Asiedu et al., 2020). Explanations detailing PGx’s uses, benefits, risks, limitations, and costs are all essential for informed consent from the consumer (Asiedu et al., 2020; Cocanour, 2017). However, consumers report having limited familiarity with both how genes can affect their response to a medication and with challenging terminology, such as “pharmacogenetics,” “metabolise,” and “enzyme,” (Allen et al., 2022; Martin et al., 2021; Meagher et al., 2022). Consumers may not be able to understand PGx testing’s impact on treatment decisions and medication management and may lack enough confidence or understanding to implement PGx-guided strategies (Doyle et al., 2023, 2025; Martin et al., 2021; Schmidlen et al., 2020; Waldman et al., 2019). These considerations are likely exacerbated for First Nations and culturally and linguistically diverse (CALD) consumers (Lewis et al., 2024; Pandey et al., 2021). Without adequate accommodations for varied traditional languages and cultural backgrounds, consumers within these communities would be unable to fully engage with decisions pertaining to their healthcare.

Additionally, consumers report preconceived ideas and misconceptions, like conflating PGx testing with disease–risk testing, such that a large proportion of participants expect receiving information regarding their cancer risk from PGx results (Dressler et al., 2019). Many people may also expect PGx testing to identify the exact medication that will provide the most optimal treatment for their condition, which may lead to feelings of disappointment, anger, and failure if results do not align with consumer expectations (Allen et al., 2022). These knowledge gaps and conceptual misunderstandings may limit consumers’ ability to provide informed consent and identify whether PGx information is being adequately incorporated into their care (Pacyna et al., 2025).

People also express several concerns about PGx information, including data security, insurance discrimination or insurance implications, scientific and technical limitations, and post-testing concerns regarding PGx either being unhelpful or not aligning with prior medication experiences (Allen et al., 2022; Gawronski et al., 2023; Moore et al., 2025; Pacyna et al., 2025). Financial barriers are also a major concern for some consumers (Allen et al., 2022; Gawronski et al., 2023; Pacyna et al., 2025). However, Australian consumers have not reported this concern as strongly as those in the United States (U.S.), possibly because they assume testing is Medicare-funded (Moore et al., 2025). This is despite the fact that only three PGx tests are currently funded by the Medicare system; the rest consumers pay for as an out-of-pocket cost in primary care (Stocker & Polasek, 2025).

Consequently, consumer education materials should aim to address and alleviate these misconceptions and concerns to improve the uptake and effectiveness of pharmacogenetic testing (Gawronski et al., 2023; Pacyna et al., 2025; Rosenman et al., 2017). Furthermore, educational materials should address the BRAN questions (what are the **B**enefits, **R**isks, and **A**lternatives, and what if I do **N**othing?) to effectively facilitate informed consent and shared decision making (Hoque, 2024). By increasing the accessibility of information, we can make science more equitable for society and involve a greater scope of people that seek to benefit from research (Shulman et al., 2020). Therefore, this study aims to investigate whether health consumer-facing online written PGx health information meets readability guidelines, utilise effective visual elements, and provide quality information on PGx treatment choices to foster shared decision making and informed consent.

## Materials and Methods

### Search Strategy

We conducted our searches in October 2024 using Google and Bing, which collectively make up the majority of the search engine market share in Australia (StatCounter, 2024). Through collaboration between authors, we generated 10 search queries based off key words present in the literature (e.g., “pharmacogenomics” and “gene testing,” see Figure 1 in the Results section for the full list of queries). For each query, we planned to scrape two pages of results with 10 results per page on both search engines, resulting in a predicted sample size of 400 webpages.

**Figure 1.**
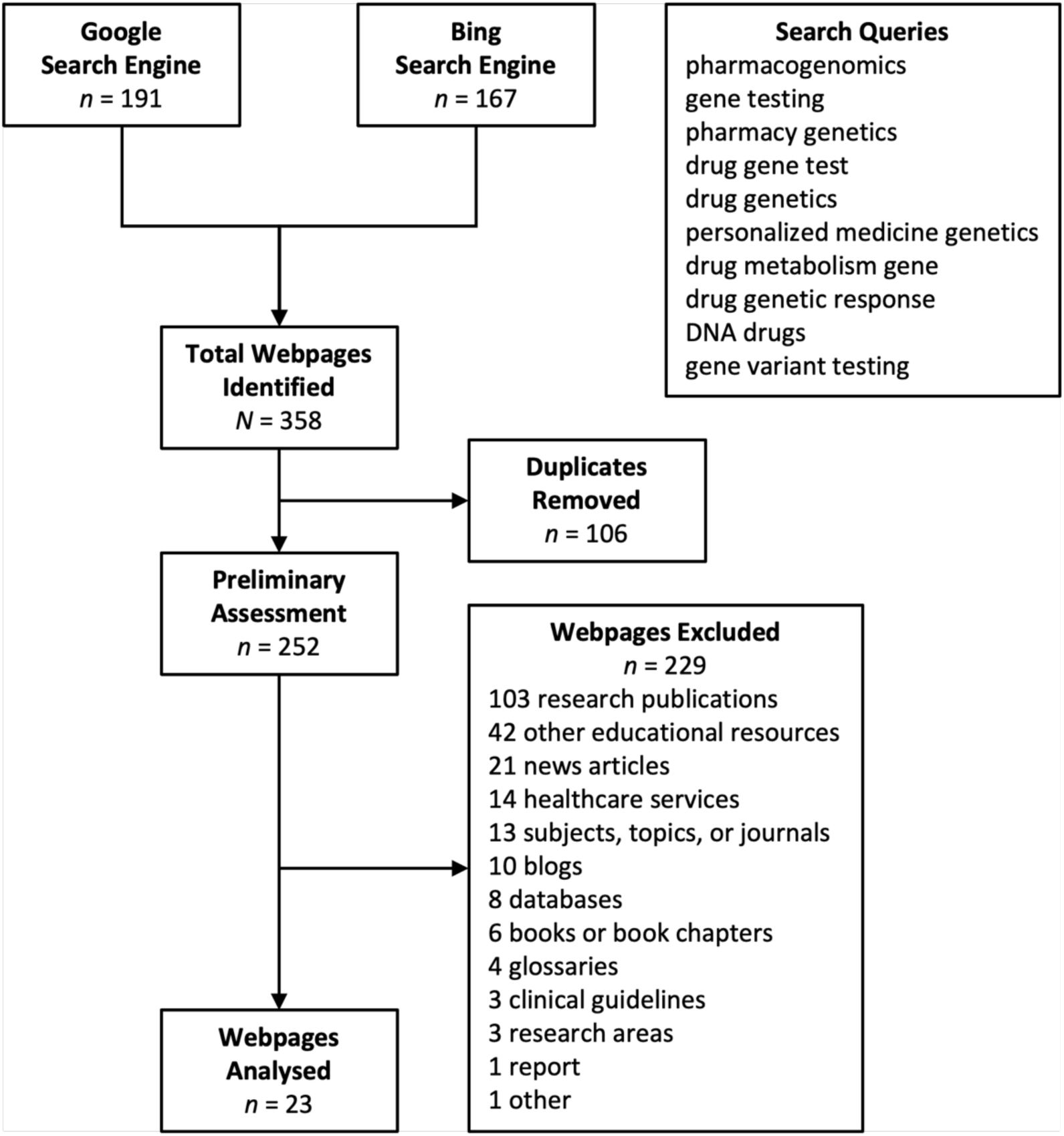
Flow Diagram of Search Engine Response Page Scrape Results.

### Inclusion and Exclusion Criteria

The primary inclusion criterion for our webpage assessment was any discussion of pharmacogenomics, drug–gene interactions, or related topics using synonymous terms, regardless of whether the discussion was extensive or limited to a single sentence. We excluded any webpages that were not written in English, were not free to access, that were not educational (for example, a medical service portal) or that exclusively targeted professionals (for example, research publications). We decided to include Wikipedia into our assessment because it is a prominent health information resource for health consumers and the general public (D. A. Smith, 2020).

## Measures

### Readability

As there is no consensus on which readability indices are best for assessing health information, we used six indices to improve the validity of the results (Badarudeen & Sabharwal, 2010). We selected the Flesch–Kincaid Grade Index (Kincaid et al., 1975), Coleman–Liau Index (Coleman & Liau, 1975), Simplified Measure of Gobbledygook Index (SMOG; McLaughlin, 1969), Gunning–Fog Index (Gunning, 1952), Automated Readability Index (ARI; E. A. Smith & Senter, 1967), and Flesch Reading Ease score (FRE; Flesch, 1948) because these readability formulae have all previously assessed readability within the medical field (Badarudeen & Sabharwal, 2010; Hutchings et al., 2024). These formulae output a score which corresponds to the necessary years of education required to comprehend the input text, with the exception of FRE, which outputs a score between 0–100, with higher scores indicating easier readability (Friedman & Hoffman-Goetz, 2006). Supplementary Table S1 shows the formula for each readability index.

### DISCERN

This study used the DISCERN questionnaire (Charnock et al., 1999) to assess the quality of consumer-facing written health information regarding pharmacogenomics. This questionnaire consists of 16 questions, scored on a 5-point scale (e.g., “Does it describe how the treatment choices affect overall quality of life?” 1 = no, 5 = yes), including items on how well the educational resources answer the BRAN questions. The full list of items is available in Supplementary Information. Scores are summed for a maximum of 80 and minimum of 16, with higher scores indicating greater quality of written consumer health information.

### Visual Elements

To assess the quality of visual elements in each webpage, we used the *TOOLKIT for Making Written Material Clear and Effective* (Centers for Medicare and Medicaid Services, 2010), a comprehensive guide to help create clear, accessible, and readable written online materials,. These elements included design choices such as page layout, fonts, text size, headings, colours, and more. Our final measure contained 32 criteria, obtained from the Toolkit Guidelines for Design sections 5.1 to 10.4. All items are available in Supplementary Information. The criteria were scored on a 3-point scale (e.g., “The material looks appealing at first glance,” −1 = No, 0 = Not applicable, 1 = Yes). Scores were summed for a maximum of 32 and minimum of −32, with higher scores indicating greater visual design quality.

### Procedure

We used a Python command line interface utilising a free trial of the Oxylabs’ *SERP Scraper API* (Oxylabs, n.d.) to scrape webpages from two search engine response pages for each search query from Google and Bing. Each webpage was screened, removing any that did not mention pharmacogenomics, pharmaceuticals or drugs and their interactions with genes, or that targeted medical professionals. Following screening, two authors (YB and MG) independently evaluated each relevant webpage and recorded their DISCERN and visual element scores in a Google Form.

To evaluate the readability of a webpage, we first extracted the relevant text while avoiding irrelevant noise, such as headers and footers, using the open-source Python command line interface Trafilatura (Version 1.12.2; Barbaresi, 2021). Trafilatura systematically extracted the relevant text from most of the webpages to text files; we manually copied the text where it failed (*n* = 5). We inspected each resulting text file to remove reference lists, as they are not part of the main body of text and so should not influence readability results. The Python module *py-readability-metrics* (DiMascio, 2020) then calculated the readability indices for each webpage. Though not formally assessed, the authors made note of any resources that mentioned the considerations affecting First Nations consumers’ access to reliable PGx testing.

### Design and Data Analysis

This study had a cross-sectional design. Analyses were performed using jamovi, version 2.6 (The jamovi project, 2025). Independent item scores from each author were first summed separately to give two total DISCERN and visual scores for each webpage and subsequently averaged to give each webpage a single DISCERN and visual score. We assessed inter-rater agreement between authors using Lin’s concordance correlation coefficient (ρc).

## Results

As shown in Figure 1, we identified 252 unique webpages from a pool of 358, of which only 23 were relevant. Table 1 shows the characteristics and descriptive statistics of the relevant webpages. Supplementary Table S2 provides the full list of these webpages, including web addresses. Most webpages originated from the U.S. and were authored by academic institutions (e.g., The Royal Australian College of General Practitioners) or government services (e.g., MedlinePlus). None of the webpages mentioned diverse consumers’ access to PGx.

**Table 1.**
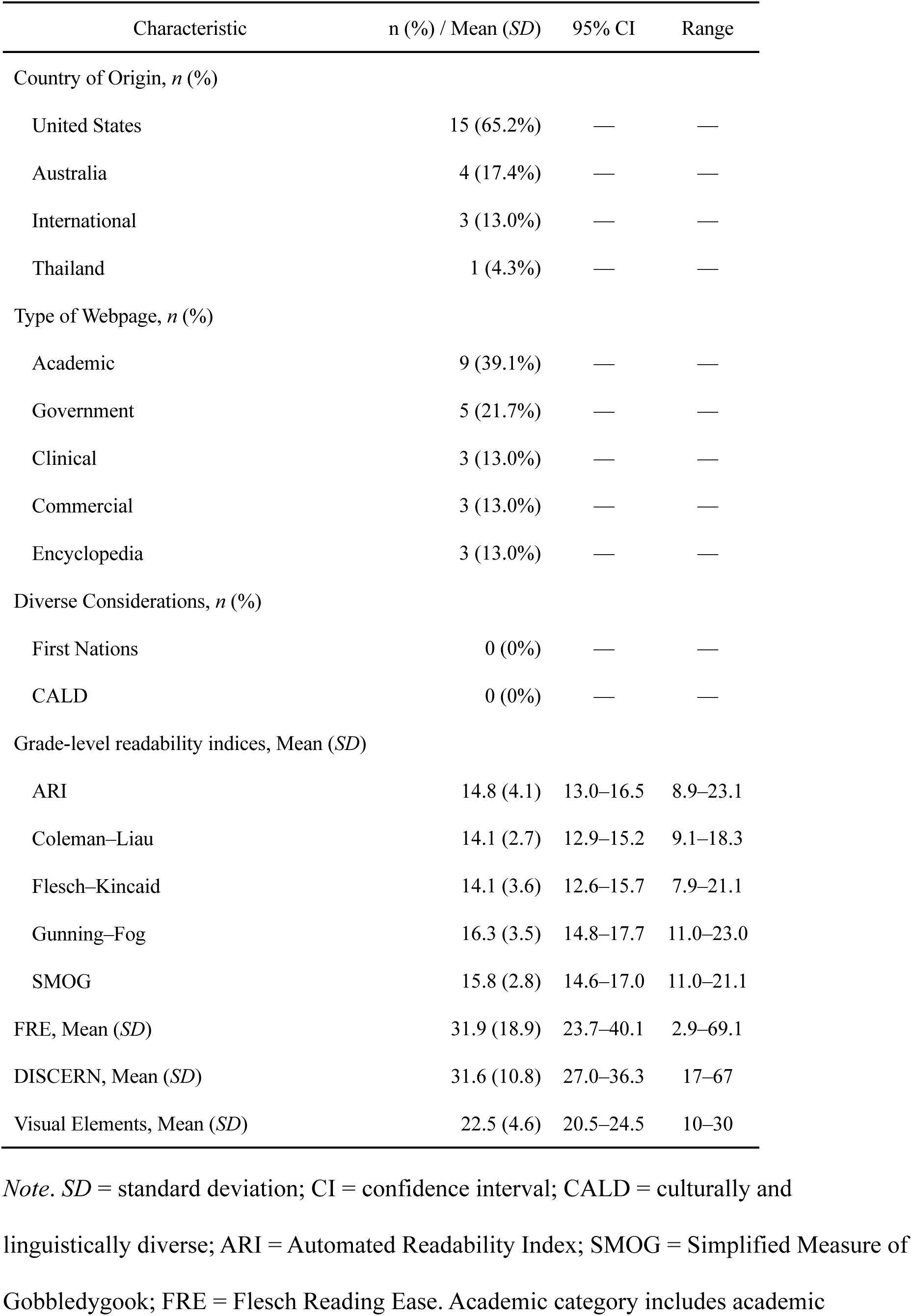
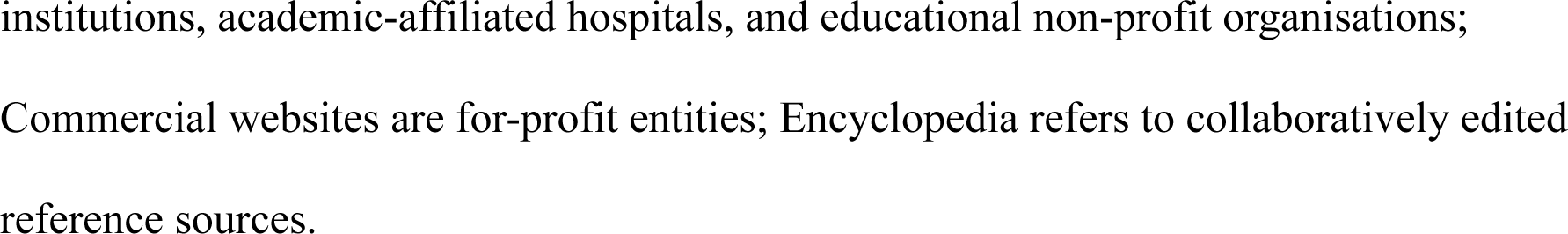
Characteristics and Descriptive Statistics of Pharmacogenomics Webpages.

| Characteristic | n (%) / Mean (SD) | 95% CI | Range |
| --- | --- | --- | --- |
| Country of Origin, *n* (%) |  |  |  |
| United States | 15 (65.2%) | — | — |
| Australia | 4 (17.4%) | — | — |
| International | 3 (13.0%) | — | — |
| Thailand | 1 (4.3%) | — | — |
| Type of Webpage, *n* (%) |  |  |  |
| Academic | 9 (39.1%) | — | — |
| Government | 5 (21.7%) | — | — |
| Clinical | 3 (13.0%) | — | — |
| Commercial | 3 (13.0%) | — | — |
| Encyclopedia | 3 (13.0%) | — | — |
| Diverse Considerations, *n* (%) |  |  |  |
| First Nations | 0 (0%) | — | — |
| CALD | 0 (0%) | — | — |
| Grade-level readability indices, Mean (SD) |  |  |  |
| ARI | 14.8 (4.1) | 13.0–16.5 | 8.9–23.1 |
| Coleman–Liau | 14.1 (2.7) | 12.9–15.2 | 9.1–18.3 |
| Flesch–Kincaid | 14.1 (3.6) | 12.6–15.7 | 7.9–21.1 |
| Gunning–Fog | 16.3 (3.5) | 14.8–17.7 | 11.0–23.0 |
| SMOG | 15.8 (2.8) | 14.6–17.0 | 11.0–21.1 |
| FRE, Mean (SD) | 31.9 (18.9) | 23.7–40.1 | 2.9–69.1 |
| DISCERN, Mean (SD) | 31.6 (10.8) | 27.0–36.3 | 17–67 |
| Visual Elements, Mean (SD) | 22.5 (4.6) | 20.5–24.5 | 10–30 |

*Note.* SD = standard deviation; CI = confidence interval; CALD = culturally and linguistically diverse; ARI = Automated Readability Index; SMOG = Simplified Measure of Gobbledygook; FRE = Flesch Reading Ease. Academic category includes academic
institutions, academic-affiliated hospitals, and educational non-profit organisations; Commercial websites are for-profit entities; Encyclopedia refers to collaboratively edited reference sources.

## Readability

Overall, the webpages had a mean (± SD) of 6,941.8 (8,204.8) letters, 1,318.6 (1,558.7) words, 57.7 (58.2) sentences, 307.8 (400.0) polysyllabic words, and an average of 1.8 (0.2) syllables per word. As shown in Table 2 below, 19 of the 23 webpages had FRE scores below 50, indicating a high reading difficulty and equating to the reading level of a university student or higher. Only two webpages (U.S. Centers for Disease Control and Prevention and MedlinePlus) were scored as standard difficulty, or 8th to 9th grade reading level. The grade-level readability indices revealed an average required reading level of 15.0 years of education (university level) and ranged between 7.9 and 23.1. As shown in Table 3 below, these indices showed very high internal consistency (Cronbach’s α = .97), with uniformly high item-rest correlations and mostly excellent Lin’s concordance coefficients (Akoglu, 2018), indicating that they ranked webpages’ readability in a highly consistent manner. Based on the Australian Government’s recommended reading level of grade 7, none of the 23 webpages were at an appropriate reading level for the general public (Australian Government, 2024). Supplementary Table S3 shows all six readability results for each webpage.

**Table 2.** Distribution of Webpages by Flesch Reading Ease Score Interpretation Categories.

| Score | Education Level | Interpretation | *n* (%) |
| --- | --- | --- | --- |
| $\geq 90$ | 5<sup>th</sup> grade | Very easy | 0 (0.0) |
| $\geq 80$ and $< 90$ | 6<sup>th</sup> grade | Easy | 0 (0.0) |
| $\geq 70$ and $< 80$ | 7<sup>th</sup> grade | Fairly easy | 0 (0.0) |
| $\geq 60$ and $< 70$ | 8<sup>th</sup> & 9<sup>th</sup> grade | Standard | 2 (8.7) |
| $\geq 50$ and $< 60$ | 10<sup>th</sup>–12<sup>th</sup> grade | Fairly difficult | 2 (8.7) |
| $\geq 30$ and $< 50$ | Undergraduate | Difficult | 10 (43.5) |
| $\geq 10$ and $< 30$ | Postgraduate | Very difficult | 5 (21.7) |
| $< 10$ | Professional | Extremely difficult | 4 (17.4) |

**Table 3.**
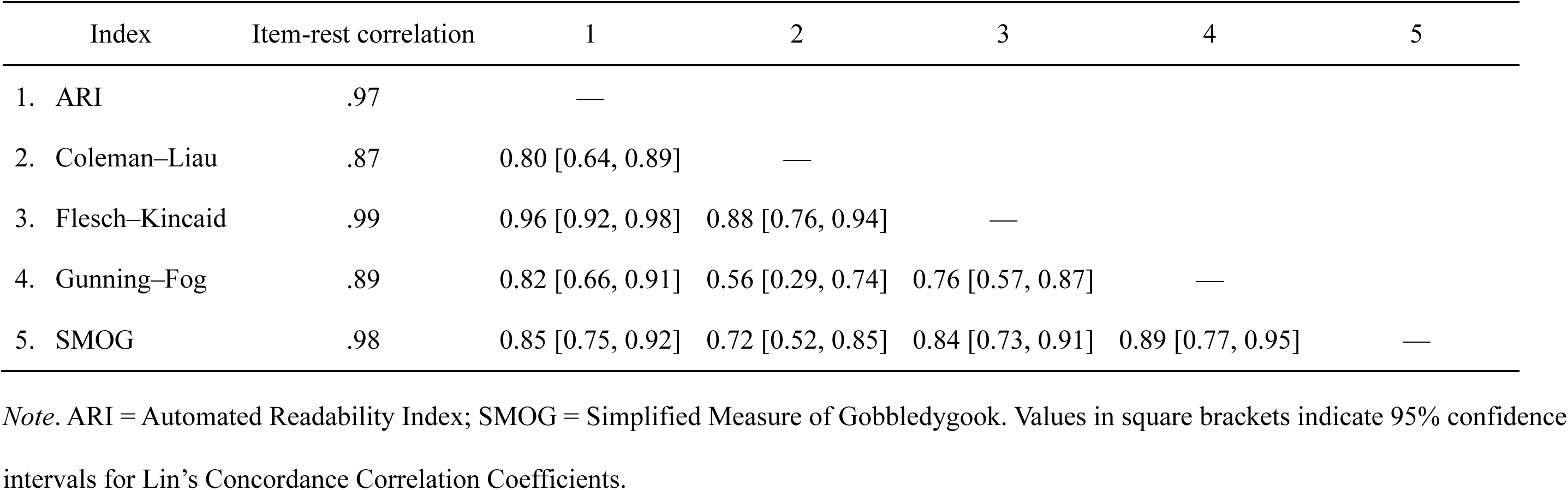
Item-Rest and Concordance Correlation Coefficients Among Readability Indices.

| Index | Item-rest correlation | 1 | 2 | 3 | 4 | 5 |
| --- | --- | --- | --- | --- | --- | --- |
| 1. ARI | .97 | — |  |  |  |  |
| 2. Coleman–Liau | .87 | 0.80 [0.64, 0.89] | — |  |  |  |
| 3. Flesch–Kincaid | .99 | 0.96 [0.92, 0.98] | 0.88 [0.76, 0.94] | — |  |  |
| 4. Gunning–Fog | .89 | 0.82 [0.66, 0.91] | 0.56 [0.29, 0.74] | 0.76 [0.57, 0.87] | — |  |
| 5. SMOG | .98 | 0.85 [0.75, 0.92] | 0.72 [0.52, 0.85] | 0.84 [0.73, 0.91] | 0.89 [0.77, 0.95] | — |

*Note.* ARI = Automated Readability Index; SMOG = Simplified Measure of Gobbledygook. Values in square brackets indicate 95% confidence intervals for Lin's Concordance Correlation Coefficients.

### DISCERN

Table 4 presents the descriptive statistics and item-rest correlations for each DISCERN item; Table 5 shows the distribution of total DISCERN scores across quality categories. Together, they reveal that the majority of webpages received a poor or very poor total DISCERN score. DISCERN had excellent internal consistency (Cronbach’s α = .95), with most items having moderate to high item-rest correlations, except for items 1 and 2, which were poor. We observed moderate overall inter-rater agreement (ρ_c_ = 0.68, 95% CI [0.42, 0.84]). Supplementary Figure S1 shows a radar chart comparing the raters’ DISCERN item scores. The items that webpages tended to achieve the most on were relevancy of the webpage, describing the benefits of treatment, and being balanced and unbiased. The items that scored the lowest were providing support for shared decision making, describing how treatment choices affect quality of life, describing the consequences of no treatment, describing alternative treatment choices, describing any associated risks, and clearly describing its aims. Many webpages did not address areas of uncertainty or the limitations of PGx and often did not report their publication date clearly, nor their sources. When webpages did provide their sources of information, they commonly referenced webpages produced by the same institution rather than providing a diverse range of alternatives. Consequently, we scored many webpages poorly on question 6 because it is difficult to identify whether a webpage is balanced and unbiased when it does not clearly provide the sources used to compile the webpage, the publication or last edited date, and diverse additional information and support. This is especially true for commercial webpages attempting to sell products, who may be biased by not disclosing the limitations, uncertainties, and risks of their treatment. Supplementary Tables S4 and S5 present the results of each DISCERN item across all webpages for both independent raters.

**Table 4.**
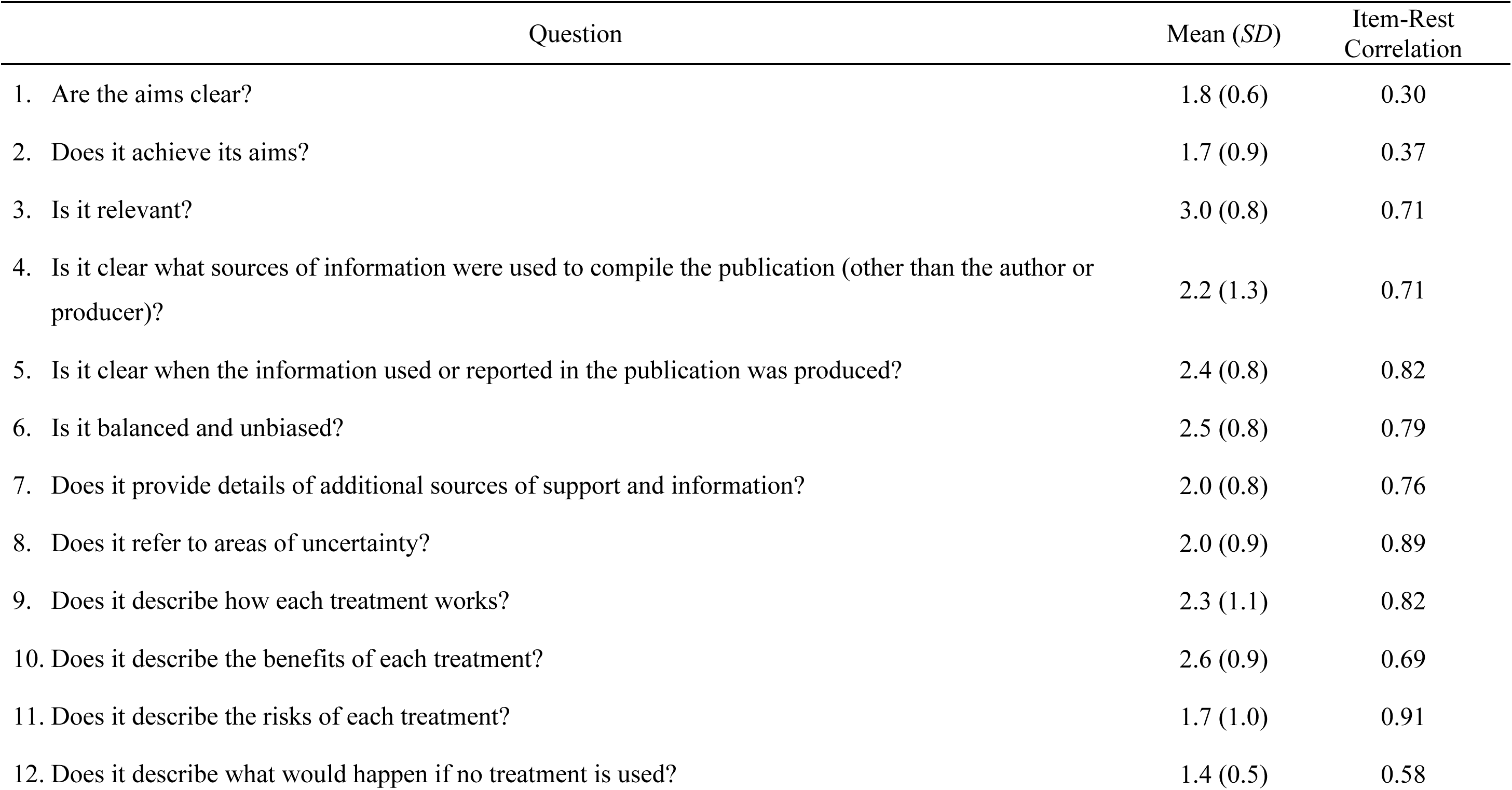
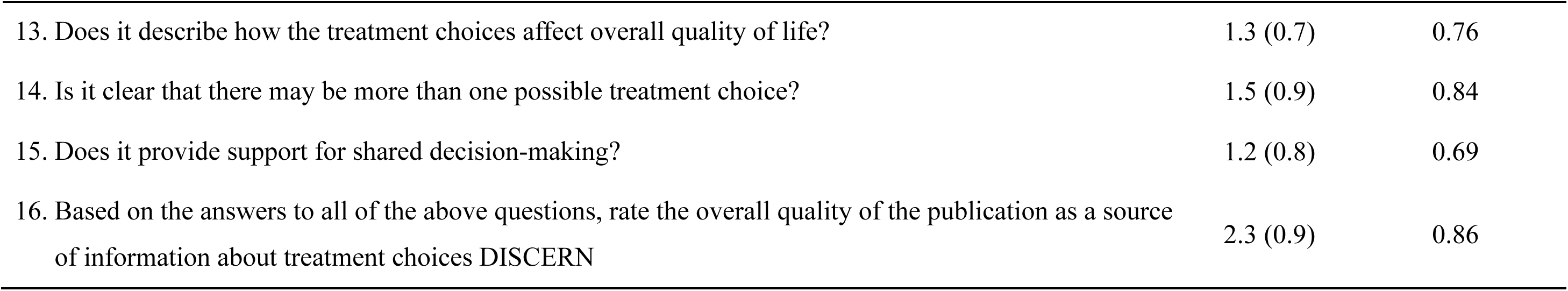
Descriptive Statistics and Item-Rest Correlations of DISCERN Items.

**Table 5.**
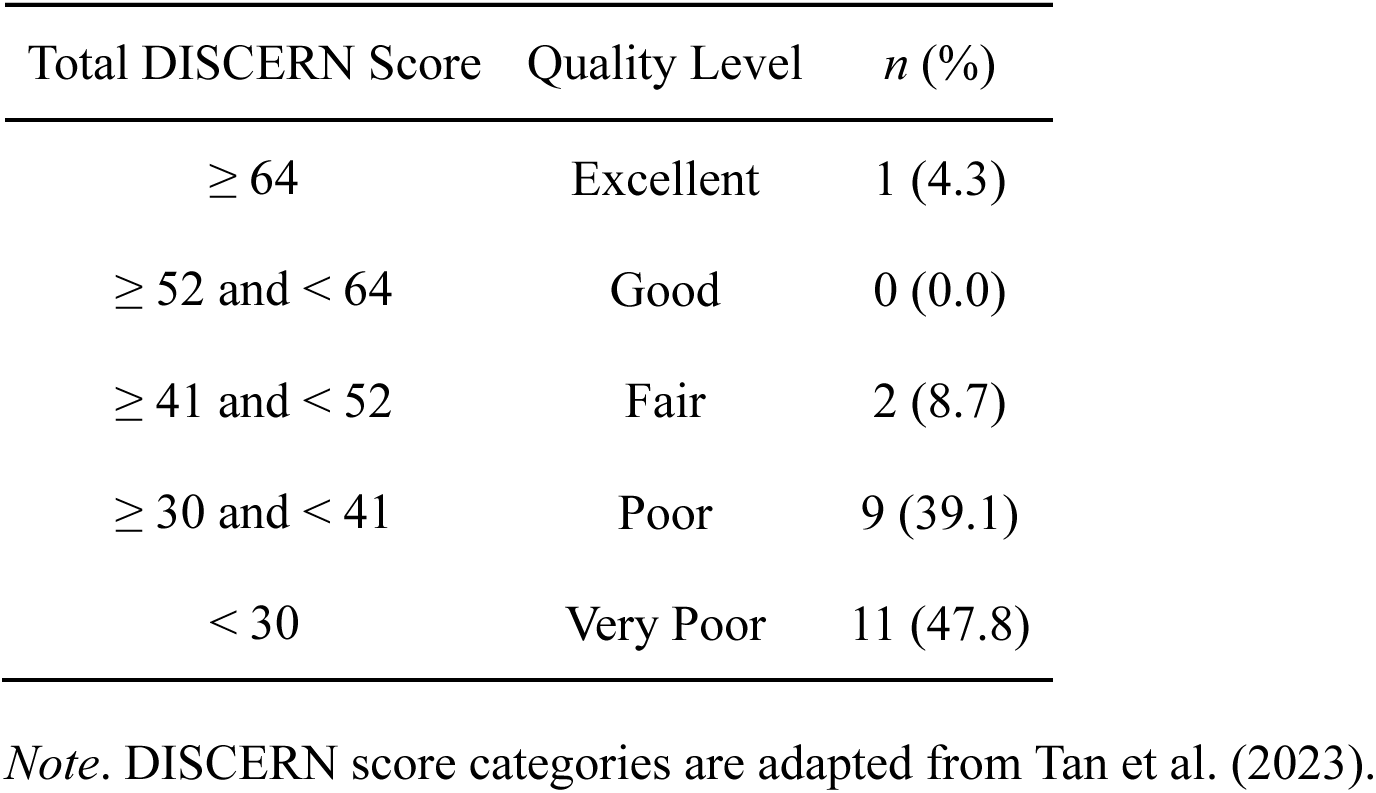
Distribution of Webpages by Total DISCERN Score Quality Categories.

### Visual Elements

Overall, the PGx health webpages fulfilled 76.9% of visual criteria, failed 6.6%, and did not apply for 16.5%. In other words, webpages successfully fulfilled on average 24.6 of the 32 criteria, failed 2.1, and did not apply for 5.3. Four webpages (17.4%) did not fail a single criterion. None of the webpages achieved the highest score because none used any calculations and so they were all marked as non-applicable for item 6.4. However, some webpages occasionally use numbers such as percentages, potentially without carefully explaining them; this may be a slight shortcoming of our scoring. Eleven of the webpages (47.8%) contained photographs, illustrations, clip art, or symbols, which mostly successfully fulfilled the relevant criteria. Eight of the webpages (34.8%) contained tables, charts, or diagrams. The criteria we scored the lowest for this section was “considerations of the likely literacy levels of the reader” because many large tables contained drug names, associated genes, and therapeutic outcomes. We felt that these tables comprised too much technical jargon which may have been overwhelming and incomprehensible to the reader. Otherwise, the least achieved criterion was the use of bold or italic text to emphasise key words and phrases, followed by the use of extra line spacing for better readability, with many webpages having dense paragraphs of text without emphasising the information most important to consumers (see Supplementary Figure S2 for an example image). We observed moderate inter-rater agreement using Lin’s concordance coefficient (ρ_c_ = 0.53, CI [0.18, 0.76]). Supplementary Tables S6 and S7 display the results of each visual element criteria across all webpages for both independent raters.

## Discussion

The primary objective of this study was to evaluate the accessibility and quality of online written PGx health information available to the Australian public. With an eight-year gap between the recommended Year 7 reading level and the average readability of the sampled webpages (university level), the available online PGx resources are largely inaccessible to the average layperson. This disparity indicates that individuals with lower literacy or health literacy are likely to struggle to understand key concepts surrounding PGx testing, its benefits, limitations, and implications for treatment decisions, despite the high prevalence and burden of adverse drug reactions in Australia and the potential for PGx to predict and reduce these risks (Bi et al., 2025; Chenchula et al., 2024; Dedefo et al., 2024; Li et al., 2021; Phillips et al., 2014; Zhang et al., 2019).

Health information that exceeds the recommended readability threshold presents a significant barrier to informed consent and shared decision making. Individuals with limited literacy may struggle to recognise and decode unfamiliar or complex words, which increases cognitive effort and reduces their ability to understand and integrate educational information, especially when materials assume prior scientific knowledge (Ames, 2019). When consumers cannot adequately comprehend health information, they may be less able to evaluate the benefits, risks, and alternatives of PGx testing, potentially undermining the informed consent process and limiting their participation in shared decision making. This is particularly relevant for PGx, where consumers must interpret complex concepts involving genetics, metabolism, and medication response (Allen et al., 2022).

Beyond readability, multimodal approaches utilising effective visual communication are essential for improving health literacy among consumers with different educational needs and preferences by simplifying information and clarifying intended messages (Moore et al., 2025; Woloshin et al., 2023). Visual aids such as diagrams, infographics, and simplified charts can help explain abstract concepts like drug–gene interactions more intuitively. Evidence suggests that including pictures in health communication materials improves consumer knowledge, particularly among individuals with lower health literacy (Schubbe et al., 2020). Although many webpages in our sample successfully incorporated professional visual design elements and occasionally included images or diagrams, these elements were not consistently used to enhance comprehension. Instead, some webpages presented dense tables of drug–gene interactions or technical terminology that may have overwhelmed readers lacking relevant background knowledge.

While the webpages in our study often succeeded in appearing professional and visually well-structured, they frequently fell short in providing substantive content required for informed consent and shared decision making. Many resources did not clearly answer all of the BRAN questions (Hoque, 2024) as they often did not describe the risks associated with PGx testing, any alternatives, or the consequences of not undertaking testing. Educational webpages that omit these elements risk presenting PGx as a purely beneficial or technical innovation. Without acknowledging uncertainties, limitations, trade-offs, or personal values and preferences, these webpages fail to adequately educate health consumers to make informed decisions about their healthcare.

To facilitate informed consent, educational webpages should also address and alleviate consumer concerns regarding privacy, insurance, and data security. Consumers frequently report uncertainty about how their genomic information will be stored, who will have access to it, and whether it could affect insurance eligibility or employment opportunities (Allen et al., 2022; Meagher et al., 2022; Moore et al., 2025). Clear explanations regarding the storage of PGx results within medical records, the chain of custody of genomic information, and data security safeguards may help mitigate these concerns. We did not observe any Australian-specific privacy or data security information in the sampled webpages.

The potential benefits of PGx are highly dependent on consumer engagement and the implementation of PGx-guided strategies in medication management (Waldman et al., 2019). Consequently, educational resources should do more to communicate how consumers’ results specifically impact their current and future medications as well as place greater focus on consumer behaviour change and clinical utility rather than just teaching genomic science (Doyle et al., 2025). Our DISCERN results highlight a lack of support for shared-decision making, suggesting that webpages may not empower consumers towards cultivating a working partnership with their healthcare providers regarding PGx testing.

## Recommendations for Organisations

Materials designed for the public must quickly and clearly convey their purpose and potential relevance to the reader in order to engage individuals with limited literacy skills (Ames, 2019). Webpages that clearly explain why PGx testing matters early, such as preventing adverse drug reactions or improving medication effectiveness, may be more effective in motivating consumers to explore the topic further and discuss it with their healthcare providers. Webpages could also include cases or personal stories to provide concrete examples of how PGx has worked for others.

Simplifying written information alone is not sufficient to achieve meaningful consumer engagement in shared decision making and informed consent, as consumers also need the necessary skills and competencies to participate in decision processes effectively (Toapanta et al., 2023). From our observations, most webpages failed to go beyond just explaining PGx. To improve this, webpages could incorporate or link to patient decision aids (Stacey et al., 2024) such as Queensland Health’s Care Companion, which facilitates collaborative treatment decision making between consumers and clinicians (Queensland Health, 2025). Additional strategies may include simply encouraging consumers to discuss PGx testing with healthcare professionals or family members, providing practical questions consumers can ask during consultations, and including prompts that encourage two-way discussion.

Another important consideration is the geographical relevance of online health information. Most webpages identified in this study originated from the U.S. and therefore may not adequately address Australian healthcare policies, Medicare funding (Stocker & Polasek, 2025), or legal protections, such as the recent legislation to ban genetic discrimination in life insurance (Australian Human Rights Commission, 2025). High-quality, accessible, understandable and *localised* educational resources are therefore necessary to ensure that consumers receive information that accurately reflects their healthcare context.

Importantly, PGx research has historically focussed on samples of European Ancestry, which affects the applicability of PGx findings across different populations (Pirmohamed, 2023). In our sample, we did not observe any mentions of the limited reliability of PGx with diverse populations. Educational webpages should acknowledge these limitations, particularly in relation to Indigenous populations, who may have unique PGx profiles distinct from major global populations (Jaya Shankar et al., 2022; Samarasinghe et al., 2023). Failure to acknowledge these considerations risks reinforcing existing health inequities by presenting PGx knowledge as universally applicable when evidence for certain populations may be limited. Educational materials must therefore also consider CALD groups as well as rural and remote communities to ensure that PGx contributes to reducing, rather than exacerbating, disparities in healthcare access and outcomes (Khatri & Assefa, 2022).

Searches for specific medications or PGx tests may lead users to commercial websites that offer testing services or related products. Such webpages may present PGx in an overly favourable manner by emphasising benefits while downplaying uncertainties, limitations, or costs. Educational webpages must transparently discuss the financial aspects of PGx testing, including potential out-of-pocket expenses and reimbursement within healthcare systems (Pacyna et al., 2025). Clear disclosure of commercial interests as well as limitations may help readers better critically evaluate information.

Developers of consumer-facing PGx resources should also consider incorporating mechanisms for user feedback and iterative improvement. Amato et al. (2025) suggest integrating mechanisms for consumer feedback to ensure ongoing refinement and improvement of consumer-facing PGx reports, ensuring they remain clear, accurate, and effective. Educational webpages could apply similar approaches by including opportunities for readers to provide feedback on clarity and usefulness. For example, webpages could incorporate prompts following the “Ask–Tell–Ask” communication framework (Fogarty & Crues, 2017), encouraging readers to reflect on their understanding with questions such as “What do you understand about pharmacogenomics?” or “Which parts of this page were difficult to understand?” Such strategies may help identify areas where information can be simplified or expanded to better meet consumer needs.

## Limitations and Future Directions

While existing research on the topic often focusses on a single domain of online resource quality (e.g., readability), our study jointly assessed readability, visual quality, and the quality of consumer-facing health information, enabling a broader evaluation of the selected resources (Patel et al., 2021; Wang et al., 2017). However, although authors practiced applying DISCERN and visual design criteria prior to data collection, moderate inter-rater agreement suggests that some subjectivity remained in scoring, particularly when assessing whether webpage aims were clearly stated. Future research should prioritise additional training and discussion of expectations between authors to improve consensus.

Furthermore, though we developed clear inclusion and exclusion criteria, our criteria allowed webpages that only mentioned PGx in a single sentence to be analysed. This potentially artificially lowered DISCERN scores as the instrument penalises webpages for not discussing treatment choices, risks, or alternatives that the webpage may not have intended to address in detail. However, it is important to note that our sample size was already quite small relative to the number of unique irrelevant webpages we identified, suggesting a notable gap of existing PGx information for patients available online. Future research should consider applying a minimum content threshold to ensure that the DISCERN tool is only used for webpages sufficiently designed as educational resources on the topic of focus. Additionally, while our study assessed informational quality, it did not evaluate webpages’ scientific accuracy or alignment with PGx clinical guidelines. This is a key oversight of our study as inaccurate content that fails to align with guidelines could have detrimental effects on consumers’ engagement with their healthcare.

This study also focused exclusively on written text and static visual design elements. We did not evaluate other forms of online health communication, such as videos, animations, or interactive infographics, which may improve accessibility for individuals with lower literacy levels. Similarly, our analysis assessed webpages primarily as they appeared on desktop devices and did not evaluate how webpage layouts adapt to mobile smartphone screens. It may be useful for future studies to evaluate resources in a variety of contexts.

Ultimately, while many PGx webpages demonstrate professional visual design, their high reading difficulty and limited support for shared decision making suggest that current online information is not optimally designed for public use. Improving readability, incorporating clearer explanations of risks and limitations, and providing tools that support consumer–clinician discussions are essential steps toward making PGx information more accessible. Without these improvements, online health information may fail to support the shift towards patient-centred personalised healthcare in Australia.

## Statements and Declarations

### Author Contributions

CRediT: Matthew J. Giblett: Conceptualisation, Data curation, Formal analysis, Investigation, Methodology, Software, Writing – original draft; Yousef Babikian: Conceptualisation, Investigation, Methodology, Writing – original draft; Dillensinh J. Jhala: Conceptualisation, Methodology, Supervision, Writing – review & editing; Sarah E. Medland: Conceptualisation, Supervision, Writing – review & editing.

### Funding Information

The work completed as part of this project and QIMR Berghofer’s Psychiatric Genetics Research Traineeship was supported by the Medical Research Future Funds grants MRF1200644, MRF2024891, and MRF2025803. S.E.M. was supported by the Australian National Health and Medical Research Council grant APP2025674.

### Declaration of Conflicting Interests

The authors declare that there are no conflicts of interest with respect to the research, authorship and/or publication of this article.

### Data Availability

The authors confirm that the data supporting the findings of this study are available within the article and its supplementary materials.

## Notes

### Competing Interest Statement

The authors have declared no competing interest.

