## Supplementary for "Investigating the Readability, Visual Design, and Quality of Online Written Pharmacogenomics Health Information for Health Consumers in Australia"

### Author Note

Matthew J. Giblett http://orcid.org/0009-0006-0400-6575

Yousef Babikian http://orcid.org/0009-0008-8942-0293

Dillensinh J. Jhala http://orcid.org/0009-0001-1101-5051

Sarah E. Medland http://orcid.org/0000-0003-1382-380X

Correspondence concerning this article should be addressed to Sarah E. Medland, Psychiatric Genetics, QIMR Berghofer, 300 Herston Road, Herston, QLD 4006, Australia; Phone: +61 7 33620101;

**DISCERN (Charnock et al., 1999)**

Scale: 1 = No (criterion is not fulfilled) to 5 = Yes (criterion is fulfilled).

1. Are the aims clear?
2. Does it achieve its aims?
3. Is it relevant?
4. Is it clear what sources of information were used to compile the publication (other than the author or producer)?
5. Is it clear when the information used or reported in the publication was produced?
6. Is it balanced and unbiased?
7. Does it provide details of additional sources of support and information?
8. Does it refer to areas of uncertainty?
9. Does it describe how each treatment works?
10. Does it describe the benefits of each treatment?
11. Does it describe the risks of each treatment?
12. Does it describe what would happen if no treatment is used?
13. Does it describe how the treatment choices affect overall quality of life?
14. Is it clear that there may be more than one possible treatment choice?
15. Does it provide support for shared decision-making?
16. Based on the answers to all of the above questions, rate the overall quality of the publication as a source of information about treatment choices DISCERN

**Visual Quality Assessment (Centers for Medicare and Medicaid Services, 2010)**

1 Overall design and page layout

1.1 The size, shape and general look of the material was designed with its purpose and users in mind.

1.2 The material looks appealing at first glance.

1.3 The material has a clear and obvious path for the eye to follow.

1.4 The material has a clear and consistent style and structure.

2 Fonts, size of print, and contrast

2.1 For the regular text, the material uses a font that is designed for ease of reading.

2.2 For headings, the material uses an easy-to-read font that contrasts with the main text.

2.3 In general, the material uses no more than 2 or 3 different typefaces.

2.4 The font size is large enough for the intended audience.

2.5 The material uses upper and lower case, not all capitals.

2.6 The material emphasises words and phrases by using italics or bold text.

2.7 For ease of reading, the material uses dark-coloured text on a very light background.

2.8 Text is not aligned sideways, on patterned or shaded background or on top of photos or other images.

2.9 For ease of reading, the material uses extra line spacing.

2.10 For ease of reading, the material uses left justification throughout.

2.11 Lines of text are an appropriate length —neither too long nor too short.

2.12 The material avoids hyphenation at the end of lines.

3 Headings, bulleted lists, and emphasising blocks of text

3.1 There is a clear hierarchy of prominent headings and sub-headings.

3.2 The material uses contrast to make the main points stand out.

3.3 Bulleted lists are well formatted.

3.4 The material effectively emphasises important blocks of text.

4 Use of colour

4.1 The material uses colours appealing to the intended readers.

4.2 The material uses colour sparingly and in a consistent and deliberate way.

4.3 The colour scheme works from a design standpoint and when printed.

4.4 The colour scheme works with diminished or limited colour perception.

5 Photographs, illustrations, clip art, and symbols

5.1 The material uses photos and illustrations that relate directly to the information to reinforce key messages.

5.2 The material uses images that are clear, uncluttered, and consistent in style.

5.3 The material uses photos and illustrations that are culturally appropriate for the intended readers.

5.4 When images include people, they are appropriate to the situation and intended audience.

6 Tables, charts, and diagrams

6.1 The material considers the likely literacy levels of the reader in the use of tables, charts, and diagrams.

6.2 Titles, headings, and other labelling is specific and clear

6.3 The material uses a clean, uncluttered layout with strong visual cues to guide the reader through the information

6.4 The material carefully explains numbers or calculations.

**Table S1**

*Readability Index Formulae*

| Index | Formula |
|---|---|
| Automated Readability (ARI; Smith & Senter, 1967) | $4.71\left(\frac{letters}{words}\right) + 0.5\left(\frac{words}{sentences}\right) - 21.43$ |
| Coleman–Liau (Coleman & Liau, 1975) | $5.88\left(\frac{letters}{words}\right) + 29.6\left(\frac{sentences}{words}\right) - 15.8$ |
| Gunning–Fog (Gunning, 1952) | $0.4\left(\left(\frac{words}{sentences}\right) + 100\left(\frac{complex\ words}{words}\right)\right)$ |
| Flesch–Kincaid (Kincaid et al., 1975) | $0.39\left(\frac{words}{sentences}\right) + 11.8\left(\frac{syllables}{words}\right) - 15.59$ |
| Flesch Reading Ease (FRE; Flesch, 1948) | $206.835 - 1.015\left(\frac{words}{sentences}\right) - 84.6\left(\frac{syllables}{words}\right)$ |
| Simplified Measure of Gobbledygook Index (SMOG; McLaughlin, 1969) | $1.043\sqrt{polysyllabic\ words \times \left(\frac{30}{sentences}\right)} + 3.1291$ |

*Note*. For the Gunning–Fog index, complex words were defined as words with three or more syllables, excluding proper nouns and compound words; for SMOG, *Polysyllabic words* are all words with three or more syllables.

**Table S2**

*Overview of Online Pharmacogenomics Health Information Resources*

| ID | Name | Source | Address | Country | Type |
|---|---|---|---|---|---|
| 1 | What is pharmacogenomics? | MedlinePlus | medlineplus.gov/genetics/understanding/genomicresearch/pharmacogenomics/ | U.S. | Government |
| 2 | What Is Pharmacogenomics (Pharmacogenetics)? | Cleveland Clinic | my.clevelandclinic.org/health/articles/pharmacogenomics | U.S. | Academic |
| 3 | Pharmacogenetic Testing \| PGx Test | Sonic Genetics | www.sonicgenetics.com.au/patient/test-information/pharmacogenomics/ | Australia | Clinical |
| 4 | Pharmacogenomics | Wikipedia | en.wikipedia.org/wiki/Pharmacogenomics | International | Encyclopedia |
| 5 | Pharmacogenomics (PGx) Testing | Genomic Diagnostics | www.genomicdiagnostics.com.au/practitioners/pharmacogenomic-testing/ | Australia | Clinical |
| 6 | RACGP - Pharmacogenomics | The Royal Australian College of General Practitioners | www.racgp.org.au/clinical-resources/clinical-guidelines/key-racgp-guidelines/view-all-racgp-guidelines/genomics-in-general-practice/genetic-tests-and-technologies/pharmacogenomics | Australia | Academic |
| 7 | Pharmacogenomics in patient care - Center for Individualized Medicine | Mayo Clinic | www.mayo.edu/research/centers-programs/center-individualized-medicine/patient-care/pharmacogenomics | U.S. | Academic |
| 8 | Pharmacogenetic Tests | MedlinePlus | medlineplus.gov/lab-tests/pharmacogenetic-tests/ | U.S. | Government |
| 9 | Genetic testing | Mayo Clinic | www.mayoclinic.org/tests-procedures/genetic-testing/about/pac-20384827 | U.S. | Academic |
| 10 | Genetic testing | Wikipedia | en.wikipedia.org/wiki/Genetic_testing | International | Encyclopedia |
| 11 | Pharmacogenetic (PGx) Testing | Australian Clinical Labs | www.clinicallabs.com.au/doctor/specialists-services/pharmacogenetic-testing/ | Australia | Clinical |
| 12 | Genetic Pharmacology | Cincinnati Children's | www.cincinnatichildrens.org/service/g/genetic-pharmacology | U.S. | Academic |
| 13 | Pharmacogenomic | Bangkok Hospital Headquarter | www.bangkokhospital.com/en/bangkok/content/pharmacogenomics | Thailand | Academic |
| 14 | Pharmacogenetics: How Genetic Testing Can Guide Medicine Decisions | Nemours KidsHealth | kidshealth.org/en/parents/pharmacogenomics.html | U.S. | Academic |
| 15 | What is the difference between precision medicine and personalized medicine? What about pharmacogenomics? | MedlinePlus | medlineplus.gov/genetics/understanding/precisionmedicine/precisionvspersonalized/ | U.S. | Government |
| 16 | What is personalized medicine? | Jackson Laboratory | www.jax.org/personalized-medicine/precision-medicine-and-you/what-is-precision-medicine | U.S. | Academic |
| 17 | Personalized medicine | Wikipedia | en.wikipedia.org/wiki/Personalized_medicine | International | Encyclopedia |
| 18 | CYP2C19 Drug Metabolism: Genetics and More | 23andMe | www.23andme.com/topics/pharmacogenetics/cyp2c19/ | U.S. | Commercial |
| 19 | CYP2D6: The Gene That Plays A Major Role In Drug Metabolism | ClarityX | clarityxdna.com/blog/learn/cyp2d6-gene-plays-major-role-drug-metabolism/ | U.S. | Commercial |
| 20 | Pharmacogenomics \| Genomics and Your Health \| CDC | U.S. Centers for Disease Control and Prevention | www.cdc.gov/genomics-and-health/about/pharmacogenetics.html | U.S. | Government |
| 21 | MTHFR Mutation Test | MedlinePlus | medlineplus.gov/lab-tests/mthfr-mutation-test/ | U.S. | Government |
| 22 | Pharmacogenomics genetic testing | Mayo Clinic | www.mayoclinichealthsystem.org/services-and-treatments/pharmacogenomics | U.S. | Academic |
| 23 | Understanding the Gene-Drug Interaction Chart | GeneSight | genesight.com/genetic-insights/understanding-the-gene-drug-interaction-chart/ | U.S. | Commercial |

*Note.* Academic category includes academic institutions, academic-affiliated hospitals, and educational non-profit organisations; Commercial websites are for-profit entities; Encyclopedia refers to collaboratively edited reference sources.

**Table S3**

*Readability of Online Pharmacogenomics Health Information Resources*

| ID | ARI | Coleman–Liau | Flesch–Kincaid | Gunning–Fog | SMOG | FRE | Letters | Words | Sentences | Polysyllabic | Average Syllables per Word |
|---|---|---|---|---|---|---|---|---|---|---|---|
| 1 | 8.9 | 10.2 | 8.7 | 10.9 | 11.4 | 59.9 | 4,119 | 869 | 54 | 114 | 1.5 |
| 2 | 13.2 | 14.4 | 13.1 | 14.8 | 15.0 | 32.6 | 6,243 | 1,153 | 63 | 274 | 1.8 |
| 3 | 11.0 | 11.8 | 11.5 | 14.3 | 14.0 | 43.1 | 4,125 | 829 | 46 | 168 | 1.7 |
| 4 | 17.0 | 15.5 | 16.4 | 18.9 | 17.9 | 19.6 | 22,973 | 4,163 | 168 | 1,121 | 1.9 |
| 5 | 16.2 | 13.1 | 14.4 | 16.8 | 15.7 | 38.3 | 1,809 | 355 | 13 | 63 | 1.7 |
| 6 | 23.1 | 17.2 | 21.1 | 23.0 | 21.1 | 3.8 | 5,823 | 1,013 | 29 | 286 | 2.0 |
| 7 | 14.7 | 16.1 | 14.0 | 14.6 | 14.7 | 26.8 | 2,337 | 410 | 22 | 90 | 1.9 |
| 8 | 14.7 | 14.2 | 13.6 | 15.9 | 15.4 | 35.2 | 1,408 | 264 | 12 | 55 | 1.8 |
| 9 | 14.6 | 13.1 | 13.6 | 17.4 | 16.1 | 38.6 | 8,307 | 1,623 | 68 | 351 | 1.7 |
| 10 | 15.7 | 13.0 | 15.3 | 18.7 | 17.3 | 30.1 | 24,550 | 4,822 | 183 | 1,124 | 1.8 |
| 11 | 17.6 | 18.3 | 17.6 | 13.4 | 18.6 | 5.0 | 5,379 | 890 | 42 | 308 | 2.1 |
| 12 | 22.5 | 18.3 | 20.4 | 21.8 | 20.4 | 2.9 | 1,884 | 316 | 10 | 91 | 2.0 |
| 13 | 20.9 | 18.1 | 19.5 | 21.8 | 20.0 | 4.8 | 4,617 | 778 | 27 | 236 | 2.0 |
| 14 | 9.3 | 9.5 | 9.8 | 11.7 | 12.1 | 56.5 | 3,366 | 736 | 40 | 99 | 1.6 |
| 15 | 11.7 | 13.4 | 12.7 | 15.7 | 15.7 | 32.5 | 1,401 | 266 | 16 | 77 | 1.9 |
| 16 | 14.3 | 14.2 | 13.8 | 17.3 | 16.0 | 32.6 | 3,964 | 742 | 35 | 177 | 1.8 |
| 17 | 16.7 | 14.0 | 16.4 | 19.7 | 18.0 | 23.2 | 32,766 | 6,227 | 233 | 1,572 | 1.8 |
| 18 | 14.5 | 14.4 | 13.1 | 15.0 | 14.5 | 37.0 | 3,869 | 719 | 34 | 134 | 1.8 |
| 19 | 13.6 | 15.5 | 14.0 | 16.8 | 15.6 | 24.0 | 5,713 | 1,017 | 59 | 283 | 2.0 |
| 20 | 9.2 | 9.1 | 8.6 | 11.4 | 11.4 | 66.0 | 4,759 | 1,058 | 56 | 116 | 1.4 |
| 21 | 9.2 | 9.7 | 7.9 | 11.1 | 11.0 | 69.1 | 5,410 | 1,172 | 66 | 124 | 1.4 |
| 22 | 18.4 | 16.5 | 17.4 | 18.6 | 17.7 | 15.4 | 893 | 157 | 6 | 39 | 1.9 |
| 23 | 11.9 | 13.4 | 12.3 | 14.2 | 14.6 | 36.1 | 3,947 | 749 | 44 | 178 | 1.8 |
| Mean | 14.7 | 14.0 | 14.1 | 16.2 | 15.8 | 31.9 | 6,941.8 | 1,318.6 | 57.7 | 307.8 | 1.8 |

**Table S4**

*Author 1 DISCERN Scores of Online Pharmacogenomics Health Information Resources*

| ID | 1 | 2 | 3 | 4 | 5 | 6 | 7 | 8 | 9 | 10 | 11 | 12 | 13 | 14 | 15 | 16 | Total |
|---|---|---|---|---|---|---|---|---|---|---|---|---|---|---|---|---|---|
| 1 | 2 | 4 | 3 | 1 | 2 | 2 | 2 | 2 | 1 | 3 | 2 | 1 | 1 | 1 | 1 | 2 | 30 |
| 2 | 1 | 0 | 4 | 4 | 5 | 5 | 2 | 3 | 4 | 4 | 3 | 2 | 1 | 2 | 1 | 4 | 45 |
| 3 | 1 | 0 | 4 | 1 | 1 | 2 | 1 | 1 | 3 | 3 | 2 | 1 | 1 | 1 | 1 | 2 | 25 |
| 4 | 1 | 0 | 2 | 3 | 4 | 4 | 1 | 2 | 3 | 2 | 1 | 1 | 1 | 1 | 1 | 1 | 28 |
| 5 | 1 | 0 | 4 | 1 | 1 | 2 | 1 | 3 | 3 | 3 | 2 | 1 | 2 | 3 | 1 | 3 | 31 |
| 6 | 1 | 0 | 3 | 3 | 2 | 4 | 2 | 2 | 1 | 3 | 1 | 1 | 1 | 1 | 1 | 3 | 29 |
| 7 | 1 | 0 | 3 | 2 | 2 | 3 | 2 | 2 | 1 | 1 | 1 | 1 | 1 | 1 | 1 | 2 | 24 |
| 8 | 1 | 0 | 2 | 3 | 2 | 3 | 2 | 2 | 1 | 2 | 1 | 1 | 1 | 2 | 1 | 3 | 27 |
| 9 | 1 | 0 | 5 | 4 | 5 | 4 | 5 | 5 | 5 | 5 | 5 | 1 | 4 | 5 | 5 | 5 | 64 |
| 10 | 1 | 0 | 3 | 5 | 4 | 4 | 1 | 2 | 3 | 1 | 1 | 1 | 1 | 2 | 1 | 3 | 33 |
| 11 | 1 | 0 | 3 | 2 | 2 | 3 | 2 | 2 | 2 | 2 | 2 | 2 | 2 | 2 | 1 | 2 | 30 |
| 12 | 1 | 0 | 3 | 1 | 1 | 2 | 2 | 2 | 1 | 3 | 1 | 1 | 1 | 1 | 1 | 2 | 23 |
| 13 | 1 | 0 | 3 | 1 | 1 | 3 | 1 | 1 | 2 | 2 | 1 | 1 | 1 | 1 | 1 | 1 | 21 |
| 14 | 1 | 0 | 3 | 2 | 3 | 4 | 2 | 2 | 3 | 2 | 1 | 1 | 1 | 2 | 1 | 3 | 31 |
| 15 | 1 | 0 | 2 | 1 | 1 | 2 | 1 | 1 | 1 | 1 | 1 | 1 | 1 | 1 | 1 | 1 | 17 |
| 16 | 1 | 0 | 3 | 1 | 1 | 2 | 1 | 1 | 1 | 2 | 1 | 1 | 1 | 1 | 1 | 2 | 20 |
| 17 | 4 | 3 | 2 | 4 | 3 | 2 | 1 | 2 | 1 | 1 | 1 | 1 | 1 | 2 | 1 | 2 | 31 |
| 18 | 3 | 3 | 3 | 3 | 2 | 2 | 2 | 3 | 2 | 2 | 2 | 1 | 1 | 2 | 1 | 3 | 35 |
| 19 | 1 | 0 | 4 | 4 | 4 | 3 | 2 | 2 | 2 | 3 | 1 | 1 | 1 | 2 | 1 | 3 | 34 |
| 20 | 1 | 0 | 4 | 2 | 3 | 4 | 4 | 3 | 3 | 2 | 2 | 2 | 2 | 1 | 1 | 3 | 37 |
| 21 | 1 | 0 | 4 | 4 | 1 | 3 | 2 | 2 | 3 | 3 | 3 | 2 | 3 | 1 | 1 | 3 | 36 |
| 22 | 1 | 0 | 2 | 1 | 1 | 2 | 1 | 1 | 1 | 1 | 1 | 1 | 1 | 1 | 1 | 1 | 17 |
| 23 | 1 | 0 | 2 | 1 | 1 | 1 | 1 | 1 | 2 | 2 | 1 | 1 | 1 | 1 | 1 | 1 | 18 |
| Mean | 1.3 | 0.4 | 3.1 | 2.3 | 2.3 | 2.9 | 1.8 | 2.0 | 2.1 | 2.3 | 1.6 | 1.2 | 1.3 | 1.6 | 1.2 | 2.4 | 29.8 |

**Table S5**

*Author 2 DISCERN Scores of Online Pharmacogenomics Health Information Resources*

| ID | 1 | 2 | 3 | 4 | 5 | 6 | 7 | 8 | 9 | 10 | 11 | 12 | 13 | 14 | 15 | 16 | Total |
|---|---|---|---|---|---|---|---|---|---|---|---|---|---|---|---|---|---|
| 1 | 2 | 3 | 3 | 1 | 2 | 2 | 2 | 2 | 1 | 2 | 1 | 2 | 1 | 1 | 1 | 1 | 27 |
| 2 | 2 | 4 | 5 | 3 | 4 | 3 | 3 | 3 | 4 | 5 | 3 | 3 | 2 | 3 | 2 | 4 | 53 |
| 3 | 1 | 0 | 3 | 1 | 1 | 2 | 1 | 2 | 1 | 2 | 1 | 1 | 1 | 1 | 1 | 2 | 21 |
| 4 | 2 | 4 | 2 | 4 | 5 | 2 | 3 | 4 | 5 | 4 | 4 | 4 | 2 | 2 | 1 | 3 | 51 |
| 5 | 2 | 3 | 3 | 1 | 2 | 2 | 2 | 1 | 2 | 4 | 1 | 2 | 1 | 1 | 2 | 3 | 32 |
| 6 | 2 | 3 | 3 | 1 | 2 | 2 | 3 | 2 | 4 | 4 | 1 | 2 | 1 | 2 | 1 | 3 | 36 |
| 7 | 2 | 2 | 2 | 1 | 1 | 1 | 1 | 3 | 2 | 2 | 1 | 1 | 1 | 1 | 1 | 2 | 24 |
| 8 | 2 | 3 | 4 | 3 | 3 | 3 | 3 | 1 | 2 | 2 | 2 | 1 | 1 | 1 | 1 | 3 | 35 |
| 9 | 3 | 4 | 5 | 3 | 5 | 4 | 4 | 5 | 4 | 5 | 5 | 2 | 4 | 5 | 5 | 4 | 67 |
| 10 | 4 | 5 | 4 | 5 | 5 | 3 | 4 | 3 | 4 | 4 | 4 | 2 | 1 | 2 | 1 | 3 | 54 |
| 11 | 2 | 3 | 3 | 1 | 2 | 2 | 1 | 1 | 1 | 1 | 1 | 1 | 1 | 1 | 1 | 1 | 23 |
| 12 | 2 | 2 | 2 | 1 | 1 | 1 | 2 | 1 | 1 | 2 | 1 | 1 | 1 | 1 | 1 | 1 | 21 |
| 13 | 2 | 3 | 3 | 1 | 1 | 2 | 1 | 1 | 2 | 4 | 1 | 1 | 1 | 1 | 1 | 2 | 27 |
| 14 | 2 | 4 | 3 | 1 | 3 | 3 | 2 | 1 | 3 | 2 | 1 | 1 | 1 | 1 | 1 | 2 | 31 |
| 15 | 2 | 2 | 1 | 1 | 2 | 1 | 3 | 1 | 1 | 1 | 1 | 1 | 1 | 1 | 1 | 1 | 21 |
| 16 | 2 | 3 | 2 | 1 | 1 | 1 | 1 | 1 | 2 | 4 | 1 | 1 | 1 | 1 | 1 | 2 | 25 |
| 17 | 3 | 4 | 3 | 4 | 5 | 2 | 2 | 3 | 4 | 4 | 4 | 2 | 2 | 1 | 1 | 3 | 47 |
| 18 | 3 | 3 | 2 | 3 | 1 | 2 | 1 | 1 | 1 | 1 | 1 | 1 | 1 | 1 | 1 | 1 | 24 |
| 19 | 3 | 4 | 2 | 2 | 4 | 2 | 2 | 1 | 2 | 3 | 1 | 2 | 1 | 1 | 1 | 2 | 33 |
| 20 | 3 | 2 | 2 | 2 | 2 | 1 | 2 | 1 | 3 | 3 | 1 | 1 | 1 | 1 | 1 | 2 | 28 |
| 21 | 2 | 3 | 4 | 3 | 3 | 2 | 3 | 2 | 4 | 3 | 2 | 2 | 1 | 1 | 1 | 3 | 39 |
| 22 | 1 | 0 | 2 | 1 | 1 | 1 | 1 | 1 | 1 | 3 | 1 | 1 | 1 | 1 | 1 | 1 | 18 |
| 23 | 3 | 4 | 3 | 1 | 1 | 3 | 2 | 2 | 3 | 2 | 1 | 2 | 1 | 1 | 1 | 2 | 32 |
| Mean | 2.3 | 3.0 | 2.9 | 2.0 | 2.5 | 2.0 | 2.1 | 1.9 | 2.5 | 2.9 | 1.7 | 1.6 | 1.3 | 1.4 | 1.3 | 2.2 | 33.4 |

**Figure S1**

*Radar Chart Comparing DISCERN Item Scores Between Authors*

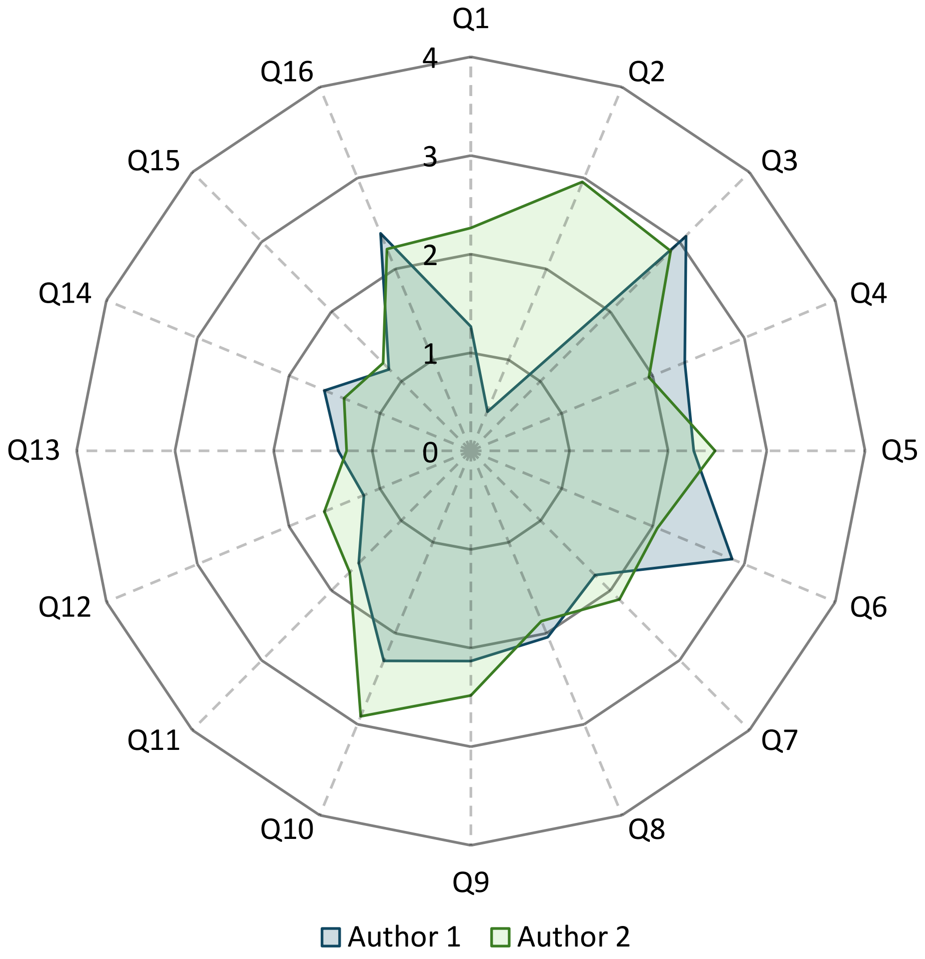

**Figure S2**

*Example of Text-Heavy Webpage Content without Typographic Emphasis Highlighting Patient-Relevant Information*

### What is pharmacogenomics?

Pharmacogenomics is the study of how genes affect a person's response to drugs. This field combines pharmacology (the science of drugs) and genomics (the study of genes and their functions) to develop effective, safe medications that can be prescribed based on a person's genetic makeup.

Many drugs that are currently available are "one size fits all," but they don't work the same way for everyone. It can be difficult to predict who will benefit from a medication, who will not respond at all, and who will experience negative side effects (called adverse drug reactions). Adverse drug reactions are a significant cause of hospitalizations and deaths in the United States.

Researchers are learning how variants in genes affect the body's response to medications. These genetic differences will be used to predict whether a medication will be effective for a particular person and which dose will help prevent adverse drug reactions. Conditions that affect a person's response to certain drugs include clopidogrel resistance, warfarin sensitivity, warfarin resistance, malignant hyperthermia, Stevens-Johnson syndrome/toxic epidermal necrolysis, and thiopurine S-methyltransferase deficiency.

The field of pharmacogenomics is growing, and new approaches are under study in clinical trials. In the future, pharmacogenomics will be used to develop tailored drugs to treat a wide range of health problems, including cardiovascular disease, Alzheimer's disease, cancer, and asthma.

**Table S6**

*Author 1 Visual Quality Assessment Scores of Online Pharmacogenomics Health Information Resources*

| ID | 1.1 | 1.2 | 1.3 | 1.4 | 2.1 | 2.2 | 2.3 | 2.4 | 2.5 | 2.6 | 2.7 | 2.8 | 2.9 | 2.10 | 2.11 | 2.12 | 3.1 | 3.2 | 3.3 | 3.4 | 4.1 | 4.2 | 4.3 | 4.4 | 5.1 | 5.2 | 5.3 | 5.4 | 6.1 | 6.2 | 6.3 | 6.4 | Total |
|---|---|---|---|---|---|---|---|---|---|---|---|---|---|---|---|---|---|---|---|---|---|---|---|---|---|---|---|---|---|---|---|---|---|
| 1 | 1 | 1 | -1 | 1 | 1 | 1 | 1 | -1 | 1 | -1 | 1 | 1 | -1 | 1 | 1 | 1 | 1 | -1 | 1 | -1 | 1 | 1 | 1 | 1 | 0 | 0 | 0 | 0 | 0 | 0 | 0 | 0 | 12 |
| 2 | 1 | 1 | 1 | 1 | 1 | 1 | 1 | 1 | 1 | 1 | 1 | 1 | 1 | 1 | 1 | 1 | 1 | 1 | 1 | 1 | 1 | 1 | 1 | 1 | 0 | 0 | 0 | 0 | 0 | 0 | 0 | 0 | 24 |
| 3 | -1 | -1 | 1 | 1 | 1 | 1 | 1 | 1 | 1 | -1 | 1 | 1 | -1 | 1 | 1 | 1 | 1 | 1 | 1 | 1 | 1 | 1 | 1 | 1 | 1 | 1 | 1 | 1 | 0 | 0 | 0 | 0 | 20 |
| 4 | 1 | -1 | 1 | 1 | 1 | 1 | 1 | 1 | 1 | 1 | 1 | 1 | -1 | 1 | -1 | 1 | 1 | 1 | 1 | 1 | -1 | 1 | 1 | 1 | 0 | 0 | 0 | 0 | -1 | 1 | -1 | 0 | 15 |
| 5 | 1 | 1 | 1 | 1 | 1 | 1 | 1 | 1 | 1 | -1 | 1 | 1 | 1 | -1 | 1 | 1 | 1 | 1 | 1 | 1 | 1 | 1 | 1 | 1 | 1 | 1 | 1 | 1 | 1 | 1 | 1 | 0 | 27 |
| 6 | -1 | -1 | 1 | 1 | 1 | 1 | 1 | 1 | 1 | -1 | 1 | 1 | -1 | 1 | 1 | 1 | 1 | 1 | 1 | -1 | 1 | 1 | 1 | -1 | 1 | 1 | 1 | 1 | 1 | 1 | 1 | 0 | 19 |
| 7 | 1 | 1 | 1 | 1 | 1 | 1 | 1 | 1 | -1 | -1 | 1 | 1 | 1 | 1 | 1 | 1 | 1 | 1 | 1 | 1 | 1 | 1 | 1 | 1 | 1 | 1 | 1 | 1 | 1 | 1 | 1 | 0 | 27 |
| 8 | 1 | 1 | 1 | 1 | 1 | 1 | 1 | 1 | 1 | -1 | 1 | 1 | -1 | 1 | 1 | 1 | 1 | 1 | 1 | -1 | 1 | 1 | 1 | 1 | 0 | 0 | 0 | 0 | 0 | 0 | 0 | 0 | 18 |
| 9 | 1 | 1 | 1 | 1 | 1 | 1 | 1 | 1 | 1 | 1 | 1 | 1 | 1 | 1 | 1 | 1 | 1 | 1 | 1 | 1 | 1 | 1 | 1 | 1 | 0 | 0 | 0 | 0 | 0 | 0 | 0 | 0 | 24 |
| 10 | -1 | -1 | 1 | 1 | 1 | 1 | 1 | -1 | 1 | 1 | 1 | 1 | -1 | 1 | 1 | 1 | 1 | 1 | 1 | 1 | -1 | -1 | -1 | 1 | 1 | 1 | 1 | 1 | 0 | 0 | 0 | 0 | 14 |
| 11 | 1 | 1 | 1 | 1 | 1 | 1 | 1 | 1 | 1 | 1 | 1 | 1 | 1 | 1 | 1 | 1 | 1 | 1 | 1 | 1 | 1 | 1 | 1 | 1 | 1 | 1 | 1 | 1 | 1 | 1 | 1 | 0 | 31 |
| 12 | 1 | 1 | 1 | 1 | 1 | 1 | 1 | 1 | 1 | 1 | 1 | 1 | 1 | 1 | 1 | 1 | 1 | 1 | 1 | 1 | 1 | 1 | 1 | 1 | 0 | 0 | 0 | 0 | 0 | 0 | 0 | 0 | 24 |
| 13 | 1 | 1 | 1 | 1 | 1 | 1 | 1 | 1 | 1 | 1 | 1 | 1 | 1 | 1 | 1 | 1 | 1 | 1 | 1 | 1 | -1 | 1 | 1 | 1 | 1 | 1 | 1 | 1 | 1 | 0 | 0 | 0 | 27 |
| 14 | 1 | 1 | 1 | 1 | 1 | 1 | 1 | 1 | 1 | 1 | 1 | 1 | 1 | 1 | 1 | 1 | 1 | 1 | 1 | 1 | 1 | 1 | 1 | 1 | 0 | 0 | 0 | 0 | 0 | 0 | 0 | 0 | 24 |
| 15 | -1 | -1 | 1 | 1 | 1 | 1 | 1 | 1 | 1 | -1 | 1 | 1 | -1 | 1 | 1 | 1 | -1 | -1 | 1 | 1 | -1 | -1 | 1 | 1 | 0 | 0 | 0 | 0 | 0 | 0 | 0 | 0 | 8 |
| 16 | 1 | 1 | 1 | 1 | 1 | 1 | 1 | 1 | 1 | 1 | 1 | 1 | 1 | 1 | 1 | 1 | 1 | 1 | 1 | 1 | 1 | 1 | 1 | 1 | 0 | 0 | 0 | 0 | 0 | 0 | 0 | 0 | 24 |
| 17 | -1 | -1 | 1 | 1 | 1 | 1 | 1 | 1 | 1 | 1 | 1 | 1 | 1 | 1 | 1 | 1 | 1 | 1 | 1 | 1 | 1 | 1 | 1 | 1 | 1 | 1 | 1 | 1 | 0 | 0 | 0 | 0 | 24 |
| 18 | 1 | 1 | 1 | 1 | 1 | 1 | 1 | 1 | 1 | 1 | 1 | 1 | 1 | 1 | 1 | 1 | 1 | 1 | 1 | 1 | 1 | 1 | 1 | 1 | 1 | 1 | 1 | 0 | 1 | 0 | 0 | 0 | 28 |
| 19 | 1 | 1 | 1 | 1 | 1 | 1 | 1 | 1 | 1 | 1 | 1 | 1 | 1 | 1 | 1 | 1 | 1 | 1 | 1 | 1 | 1 | 1 | 1 | 1 | 1 | 1 | 1 | 1 | 1 | 0 | 0 | 0 | 29 |
| 20 | 1 | 1 | 1 | 1 | 1 | 1 | 1 | 1 | 1 | 1 | 1 | 1 | 1 | 1 | 1 | 1 | 1 | 1 | 0 | 1 | 1 | 1 | 1 | 1 | 1 | 1 | 1 | 1 | 1 | 1 | 1 | 0 | 30 |
| 21 | 1 | 1 | 1 | 1 | 1 | 1 | 1 | 1 | 1 | 1 | 1 | 1 | 1 | 1 | 1 | 1 | 1 | 1 | 1 | 1 | 1 | 1 | 1 | 1 | 0 | 0 | 0 | 0 | 0 | 0 | 0 | 0 | 24 |
| 22 | 1 | 1 | 1 | 1 | 1 | 1 | 1 | 1 | 1 | 1 | 1 | 1 | 1 | 1 | 1 | 1 | 1 | 1 | 1 | 1 | 1 | 1 | 1 | 1 | 0 | 0 | 0 | 0 | 0 | 0 | 0 | 0 | 24 |
| 23 | 1 | 1 | 1 | 1 | 1 | 1 | 1 | 1 | 1 | 1 | 1 | 1 | 1 | 1 | 1 | 1 | 1 | 1 | 1 | 1 | 1 | 1 | 1 | 1 | 0 | 0 | 0 | 0 | 0 | 0 | 0 | 0 | 24 |
| % Achieved | 78 | 74 | 96 | 100 | 100 | 100 | 100 | 91 | 96 | 70 | 100 | 100 | 70 | 96 | 96 | 100 | 96 | 91 | 96 | 87 | 83 | 91 | 96 | 96 | 48 | 48 | 48 | 43 | 35 | 26 | 22 | 0 | 77.2 |
| % Failed | 22 | 26 | 4 | 0 | 0 | 0 | 0 | 9 | 4 | 30 | 0 | 0 | 30 | 4 | 4 | 0 | 4 | 9 | 0 | 13 | 17 | 9 | 4 | 4 | 0 | 0 | 0 | 0 | 4 | 0 | 4 | 0 | 6.4 |

**Table S7**

*Author 2 Visual Quality Assessment Scores of Online Pharmacogenomics Health Information Resources*

| ID | 1.1 | 1.2 | 1.3 | 1.4 | 2.1 | 2.2 | 2.3 | 2.4 | 2.5 | 2.6 | 2.7 | 2.8 | 2.9 | 2.10 | 2.11 | 2.12 | 3.1 | 3.2 | 3.3 | 3.4 | 4.1 | 4.2 | 4.3 | 4.4 | 5.1 | 5.2 | 5.3 | 5.4 | 6.1 | 6.2 | 6.3 | 6.4 | Total |
|---|---|---|---|---|---|---|---|---|---|---|---|---|---|---|---|---|---|---|---|---|---|---|---|---|---|---|---|---|---|---|---|---|---|
| 1 | -1 | 1 | 1 | 1 | 1 | 1 | 1 | -1 | 1 | -1 | 1 | 1 | 1 | 1 | 1 | 1 | 1 | -1 | 1 | 1 | 1 | 1 | 1 | 1 | 0 | 0 | 0 | 0 | 0 | 0 | 0 | 0 | 16 |
| 2 | 1 | 1 | 1 | 1 | 1 | 1 | 1 | 1 | 1 | 1 | 1 | 1 | 1 | 1 | 1 | 1 | 1 | 1 | 1 | 1 | 1 | 1 | 1 | 1 | 0 | 0 | 0 | 0 | 0 | 0 | 0 | 0 | 24 |
| 3 | 1 | 1 | 1 | 1 | 1 | 1 | 1 | 1 | 1 | -1 | 1 | 1 | 1 | 1 | 1 | 1 | 1 | 1 | 1 | 1 | 1 | 1 | 1 | 1 | 1 | 1 | 1 | 1 | 0 | 0 | 0 | 0 | 26 |
| 4 | -1 | -1 | 1 | 1 | 1 | 1 | 1 | 1 | 1 | 1 | 1 | 1 | -1 | 1 | 1 | 1 | 1 | 1 | 1 | 1 | 1 | 1 | 1 | 1 | 0 | 0 | 0 | 0 | -1 | 1 | -1 | 0 | 17 |
| 5 | 1 | 1 | 1 | 1 | 1 | 1 | 1 | 1 | 1 | 1 | 1 | 1 | 1 | 1 | 1 | 1 | 1 | 1 | 1 | 1 | 1 | 1 | 1 | -1 | 1 | 1 | 1 | 1 | 1 | 1 | 1 | 0 | 29 |
| 6 | 1 | 1 | 1 | 1 | 1 | 1 | 1 | 1 | 1 | 1 | 1 | 1 | 1 | 1 | 1 | 1 | 1 | 1 | 1 | 1 | 1 | 1 | 1 | 1 | 1 | 1 | 1 | 1 | -1 | 1 | 1 | 0 | 29 |
| 7 | 1 | 1 | 1 | 1 | 1 | 1 | 1 | 1 | -1 | -1 | 1 | 1 | -1 | 1 | 1 | 1 | 1 | 1 | 1 | 1 | 1 | 1 | 1 | 1 | 1 | 1 | 1 | 1 | 0 | 0 | 0 | 0 | 22 |
| 8 | 1 | 1 | 1 | 1 | 1 | 1 | 1 | -1 | 1 | 1 | 1 | 1 | 1 | 1 | 1 | 1 | 1 | 1 | 1 | 1 | 1 | 1 | 1 | 1 | 0 | 0 | 0 | 0 | 0 | 0 | 0 | 0 | 22 |
| 9 | 1 | 1 | 1 | 1 | 1 | 1 | 1 | 1 | 1 | 1 | 1 | 1 | 1 | 1 | 1 | 1 | 1 | 1 | 1 | 1 | 1 | 1 | 1 | 1 | 0 | 0 | 0 | 0 | 0 | 0 | 0 | 0 | 24 |
| 10 | 1 | 1 | 1 | 1 | 1 | 1 | 1 | 1 | 1 | 1 | 1 | 1 | -1 | 1 | 1 | 1 | 1 | 1 | 1 | 1 | 1 | 1 | 1 | -1 | 1 | 1 | 1 | 1 | 0 | 0 | 0 | 0 | 24 |
| 11 | 1 | 1 | 1 | 1 | 1 | 1 | 1 | 1 | 1 | 1 | 1 | 1 | 1 | 1 | 1 | 1 | 1 | 1 | 1 | 1 | 1 | 1 | 1 | 1 | -1 | 1 | 1 | 1 | -1 | 1 | 1 | 0 | 27 |
| 12 | 1 | 1 | 1 | 1 | 1 | 1 | 1 | 1 | 1 | 1 | 1 | 1 | -1 | 1 | 1 | 1 | 1 | 1 | 1 | 1 | 1 | 1 | 1 | 1 | 0 | 0 | 0 | 0 | 0 | 0 | 0 | 0 | 22 |
| 13 | 1 | 1 | 1 | 1 | 1 | 1 | 1 | -1 | 1 | 1 | 1 | 1 | 1 | 1 | 1 | 1 | 1 | 1 | 1 | 1 | 1 | 1 | 1 | 1 | 1 | 1 | 1 | 1 | 1 | 0 | 0 | 0 | 27 |
| 14 | 1 | 1 | 1 | 1 | 1 | 1 | 1 | 1 | 1 | 1 | 1 | 1 | 1 | 1 | 1 | 1 | 1 | 1 | 1 | 1 | 1 | 1 | 1 | 1 | 0 | 0 | 0 | 0 | 0 | 0 | 0 | 0 | 24 |
| 15 | 1 | 1 | 1 | 1 | 1 | 1 | 1 | -1 | 1 | -1 | 1 | 1 | -1 | 1 | 1 | 1 | -1 | -1 | 1 | 1 | 1 | 1 | 1 | -1 | 0 | 0 | 0 | 0 | 0 | 0 | 0 | 0 | 12 |
| 16 | 1 | 1 | 1 | 1 | 1 | 1 | 1 | 1 | 1 | -1 | 1 | 1 | 1 | 1 | 1 | 1 | 1 | -1 | 1 | -1 | 1 | 1 | 1 | 1 | 0 | 0 | 0 | 0 | 0 | 0 | 0 | 0 | 18 |
| 17 | 1 | -1 | 1 | 1 | 1 | 1 | 1 | 1 | 1 | 1 | 1 | 1 | -1 | 1 | 1 | 1 | 1 | 1 | 1 | 1 | 1 | 1 | 1 | -1 | 1 | 1 | 1 | 1 | 0 | 0 | 0 | 0 | 22 |
| 18 | 1 | 1 | 1 | 1 | 1 | 1 | 1 | 1 | 1 | -1 | 1 | 1 | 1 | 1 | 1 | 1 | 1 | -1 | 1 | -1 | 1 | 1 | 1 | 1 | 1 | 1 | 1 | 0 | -1 | 0 | 0 | 0 | 20 |
| 19 | 1 | 1 | 1 | 1 | 1 | 1 | 1 | -1 | 1 | -1 | 1 | 1 | 1 | 1 | 1 | 1 | 1 | 1 | 1 | -1 | 1 | 1 | 1 | 1 | 1 | -1 | 1 | 1 | 1 | 0 | 0 | 0 | 21 |
| 20 | 1 | 1 | 1 | 1 | 1 | 1 | 1 | 1 | 1 | 1 | 1 | 1 | 1 | 1 | 1 | 1 | 1 | 1 | 0 | 1 | 1 | 1 | 1 | 1 | 1 | 1 | 1 | 1 | 1 | 1 | 1 | 0 | 30 |
| 21 | 1 | 1 | 1 | 1 | 1 | 1 | 1 | -1 | 1 | 1 | 1 | 1 | 1 | 1 | 1 | 1 | 1 | 1 | 1 | 1 | 1 | 1 | 1 | -1 | 0 | 0 | 0 | 0 | 0 | 0 | 0 | 0 | 20 |
| 22 | 1 | 1 | 1 | 1 | 1 | 1 | 1 | 1 | 1 | -1 | 1 | 1 | 1 | 1 | 1 | 1 | 1 | 1 | 1 | -1 | 1 | 1 | 1 | 1 | 0 | 0 | 0 | 0 | 0 | 0 | 0 | 0 | 20 |
| 23 | 1 | 1 | 1 | 1 | 1 | 1 | 1 | 1 | 1 | -1 | 1 | 1 | 1 | 1 | -1 | 1 | 1 | 1 | 1 | -1 | 1 | 1 | 1 | 1 | 0 | 0 | 0 | 0 | 0 | 0 | 0 | 0 | 18 |
| % Achieved | 91 | 91 | 100 | 100 | 100 | 100 | 100 | 74 | 96 | 61 | 100 | 100 | 74 | 100 | 96 | 100 | 96 | 83 | 96 | 78 | 100 | 100 | 100 | 78 | 43 | 43 | 48 | 43 | 17 | 22 | 17 | 0 | 76.5 |
| % Failed | 9 | 9 | 0 | 0 | 0 | 0 | 0 | 26 | 4 | 39 | 0 | 0 | 26 | 0 | 4 | 0 | 4 | 17 | 0 | 22 | 0 | 0 | 0 | 22 | 4 | 4 | 0 | 0 | 17 | 0 | 4 | 0 | 6.7 |
